# Persistence with oral bisphosphonates and denosumab among older adults in primary care in Ireland

**DOI:** 10.1101/2020.10.05.20207290

**Authors:** Mary E. Walsh, Tom Fahey, Frank Moriarty

## Abstract

**Purpose:** Gaps in pharmacological treatment for osteoporosis can reduce effectiveness. This study aimed to estimate persistence rates for oral bisphosphonates and denosumab in older primary care patients and identify factors associated with discontinuation.

**Methods:** Older patients newly prescribed oral bisphosphonates or denosumab between 2012 and 2017 were identified from 44 general practices (GP) in Ireland. Persistence without a coverage gap of >90 days was calculated for both medications from therapy initiation. Factors associated with time to discontinuation were explored using Cox regression analysis. Exposures included age-group, osteoporosis diagnosis, fracture history, calcium/vitamin D prescription, number of other medications, health cover, dosing frequency (bisphosphonates) and previous bone-health medication (denosumab).

**Results:** Of 41,901 patients, n=1,569 newly initiated on oral bisphosphonates and n=1,615 on denosumab. Two-year persistence was 49.4% for oral bisphosphonates and 53.8% for denosumab and <10% were switched to other medication. Having state-funded health cover was associated with a lower hazard of discontinuation for both oral bisphosphonates (HR=0.49, 95%CI=0.36-0.66, p<0.01) and denosumab (HR=0.71, 95%CI=0.57-0.89, p<0.01). Older age-group, number of medications and calcium/vitamin D prescription were also associated with better bisphosphonate persistence while having osteoporosis diagnosed was associated with better denosumab persistence.

**Conclusion:** Persistence for osteoporosis medications is sub-optimal. Of concern, few patients are switched to other bone-health treatments when denosumab is stopped which could increase fracture risk. Free access to GP services and medications may have resulted in better medication persistence in this cohort. Future research should explore prescribing choices in primary-care osteoporosis management and evaluate cost-effectiveness of interventions for improving persistence.

**SUMMARY:** Gaps in pharmacological treatment for osteoporosis can reduce its effectiveness. This study found approximately half of older adults in primary care newly initiated on bisphosphonates or denosumab were still taking these after 2 years. Abrupt discontinuation of denosumab without switching to an alternative is concerning due to increased fracture risk.

## INTRODUCTION

Osteoporosis and associated fragility fractures can result in significant disability, morbidity and mortality with 20% of individuals who experience a hip fracture dying in the first year [1, 2]. It is estimated that one in five women and one in twenty men over the age of 60 have osteoporosis and 3% of older adults are expected to experience fragility fractures annually [3, 4]. Adults at high fracture risk, including those with osteoporosis, previous fractures or those who take medication that reduces bone quality, should be offered pharmacological treatment where no contraindication exists [5-8]

Oral bisphosphonates have been shown to prevent fractures in men and women and they are the most cost-effective initial therapy for osteoporosis [6, 8, 9]. Denosumab, a newer antiresorptive treatment, involves six-monthly administration by subcutaneous injection. It is recommended in patients with high fracture-risk where they are unable to take oral bisphosphonates due to difficulties with administration or intolerance caused by upper gastrointestinal symptoms [6-8]. Denosumab has been shown to prevent fractures in women [7]. While research in men remains limited, it improves bone mineral density (BMD) and has shown an effect on fracture incidence in particular cohorts [7, 9, 10]. To be cost-effective and result in optimal fracture reduction, it is important that oral bisphosphonates and denosumab are prescribed and taken correctly without unwarranted gaps in treatment or early cessation [11, 12]. Adherence (the extent to which a patient acts in accordance with the prescribed interval and dose regimen) and persistence (the accumulation of time from initiation to discontinuation of therapy) have both been found to be suboptimal in oral bisphosphonate and denosumab use [13, 14].

Clinical guidelines recommend that bisphosphonates are continued without a break for a period of at least three years and for up to ten years in those deemed to be at high risk of fracture [6, 15-17]. However, a large recently published systematic review found that 2-year persistence for oral bisphosphonates was less than 30% in most studies and that only 35% to 48% of patients are adherent at 2 years [13]. Persistence in denosumab treatment is particularly important, as the suppression of bone resorption rapidly reverses where treatment is delayed by as little as three months [18, 19]. There is some evidence that this could result in rebound vertebral fractures [20]. A recent systematic review including 16 studies of denosumab showed that average 2-year persistence was only 55% [14]. Treatment with oral bisphosphonates after stopping denosumab is protective against negative effects in most patients after one year of treatment, however stronger replacement treatments may be required for patients taking denosumab for longer periods [21, 22].

Internationally, General Practitioners (GPs) have reported uncertainty about prescription breaks in bisphosphonate treatment and cessation of denosumab [23]. Recent estimates of persistence for these medications are not available in primary care in Ireland and so the extent of the problem in the Irish setting is unknown. Furthermore, identification of factors associated with early discontinuation of these medications in a large representative primary care database could reveal circumstances in which education or input from specialists would be warranted.

### Study objectives

The aim of this study is to estimate persistence rates for oral bisphosphonates and denosumab in a cohort of older primary care patients in Ireland who are newly prescribed these medications and to identify factors associated with time to discontinuation.

## METHODS

### Study Design

The REporting of studies Conducted using Observational Routinely-collected health Data (RECORD) statement was used in the conduct and reporting of this retrospective cohort study.[24]

### Setting and data source

Data were collected as part of a larger study from 44 general practices in the Republic of Ireland in the areas of Dublin (n=30), Galway (n=11), and Cork (n=3) using the patient management software Socrates (www.socrates.ie) between January 2011 and 2017 [25]. Data included demographic, clinical, prescribing and hospitalisation records of patients who were 65 years and older at the date of data extraction (2017). Ethical approval was obtained from the Irish College of General Practitioners.

### Participants

Patients were eligible for inclusion in analysis if they were newly prescribed oral bisphosphonates or denosumab during the study period. Cohorts of patients initiated on oral bisphosphonates and denosumab were defined and analysed separately, resulting in potential overlap between these groups.

Prescriptions were identified from two sources: GP prescription records (WHO Anatomical Therapeutic Chemical classification codes) and discharge summaries of hospitalisation records (based on free-text trade and generic names). See Online Resource 1 for detailed search terms and codes.

The start of the observation period for each individual was defined as their first recorded GP consultation, prescription or hospitalisation within the dataset. For the oral bisphosphonate cohort, as it is a first-line treatment, they were defined as “newly prescribed” if they received a first prescription for a bisphosphonate and had at least 12 months of observation before this without receiving any bone-health medication prescription (see Online Resource 1 for definition and codes). For the denosumab cohort, they were defined as “newly prescribed” if they received a first denosumab prescription and had at least 12 months of observation before this without a denosumab prescription.

### Estimate of persistence

Persistence was defined as the time from initiation to discontinuation of therapy [13]. Discontinuation was considered to have occurred if there was a gap in coverage of prescriptions of more than 90 days. The coverage of prescriptions for oral bisphosphonates was calculated based on specified duration and number of issues detailed in GP prescription records, while each prescription of denosumab covered a six-month period (168 days). For the small proportion of prescriptions that were based on hospital discharge summaries, a six-month prescription (168 days) was assumed. All patients were observed for as long as data allowed after initiation of medication. For calculation of two-year persistence, patients were excluded if the initiation of medication occurred less than two-years before the end of the data-collection period. The number of patients who switched to an alternative bone-health medication within 90 days of the end of coverage period of the initial medication was calculated. These patients were subsequently excluded from the estimate of 2-year persistence.

#### Statistical analysis

Demographic and clinical variables were described for bisphosphonate and denosumab cohorts. Two-year persistence for bisphosphonates and denosumab was calculated with 95% confidence intervals.

### Factors associated with time to discontinuation

Time to discontinuation of medication was calculated in days for oral bisphosphonates and for denosumab for each patient who had at least 12 months of data after medication initiation. Patients who were found to switch to an alternative bone-health medication were excluded from time to discontinuation analysis.

#### Exposures

Exposures were defined during the time-period prior to medication initiation. These included age at the point of medication initiation, a record of osteoporosis, fragility fracture or calcium/ vitamin prescription in GP or hospitalisation records (Online Resource 1), number of unique prescribed medications in the 12 months prior to initiation and health cover type. Number of medications was analysed categorically (0-5, 6-10, 11-15 and >15 medications). Health cover type was grouped into three categories relevant to the Irish healthcare system based on whether patients are required to pay at the point of care: “General Medical Services scheme” (GMS, covering GP care, hospital care and medications), Doctor Visit Card (DVC, covering GP care only), and Private [26]. For oral bisphosphonates, dosing frequency of medication (weekly or monthly) was also included as an exposure. For the denosumab cohort, whether the patient had been on previous bone-health medication was included in the analysis.

#### Statistical analysis

Separate Kaplan Meier curves were generated to explore time to discontinuation of oral bisphosphonates (by dosing frequency) and denosumab. Factors associated with time to discontinuation were explored using univariable and multivariable Cox regression for oral bisphosphonates and denosumab. Unadjusted and adjusted Hazard Ratios (HRs) were calculated with 95% confidence intervals (CIs). Confidence intervals were adjusted for clustering of patients within GP practices. Stata 16 (StataCorp. 2019) was used for analyses and statistical significance was assumed at p<0.05.

## RESULTS

### Participants

Figure 1 shows a flow-diagram of selected patients. From 41,901 patients, n=1,569 newly initiated on oral bisphosphonates and n=1,615 on denosumab. Characteristics of the cohorts are presented in Table 1. The majority of prescriptions were identified from GP records rather than hospital discharge summaries. In the bisphosphonate cohort, 89% (n=1,391) were prescribed a medication with a weekly regimen, while 11% (n=178) were prescribed monthly dosing frequencies. In the denosumab cohort, n=689 individuals (43% of those who initiated) had been prescribed a different bone-health medication previously, while n=926 (57%) were observed to initiate directly onto denosumab. In total, 56% and 51% of the bisphosphonate and denosumab cohorts were observed to discontinue the medication. Only 9% and 6% of those who discontinued bisphosphonates and denosumab respectively were switched to a different bone-health medication within 90 days of the end of the coverage period (Table 1).

**Table 1.** Cohort characteristics.

| **Table 1. Cohort characteristics** |  |  |
| --- | --- | --- |
|  | **Bisphosphonates cohort**<br>**(n=1,569)** | **Denosumab cohort**<br>**(n=1,615)** |
| Age (mean (SD)) | 76.6 (SD=8.1), | 78.4 (SD=8.2) |
| Female sex (n (%)) | 1,242 (79.3%) | 1,463 (90.6%) |
| *Health Cover:* |  |  |
| Private and Other | 270 (17.2%) | 263 (16.3%) |
| GMS | 1,156 (73.7%) | 1,157 (71.6%) |
| DVC | 142 (9.1%) | 195 (12.1%) |
| Osteoporosis diagnosis | 470 (30.0%) | 767 (47.5%) |
| Fracture history | 157 (10.0%) | 185 (11.5%) |
| Calcium/ vitamin D prescription | 1,294 (82.5%) | 1,415 (87.6%) |
| *Source of prescription:* |  |  |
| Hospitalisation records | 178 (11.3%) | 75 (4.6%) |
| General practice prescriptions | 1391 (88.7%) | 1540 (95.4%) |
| Months between medication initiation and end of data-collection period (mean (SD)) | 41.4 (SD=16.2) | 36.9 (SD=15.6) |
| Discontinued medication (n (%)) | 882 (56.3%) | 829 (51.3%) |
| Switched to alternative bone-health medication (% of discontinued): | 81 (9.2%) | 47 (5.7%) |
| *Replacement medication:* |  |  |
| Oral Bisphosphonates (n) | N/A | 39 |
| Denosumab (n) | 78 | N/A |
| Raloxifene (n) | 0 | 2 |
| Parathyroid hormone (n) | 0 | 3 |
| Strontium (n) | 3 | 3 |

**Figure 1.**
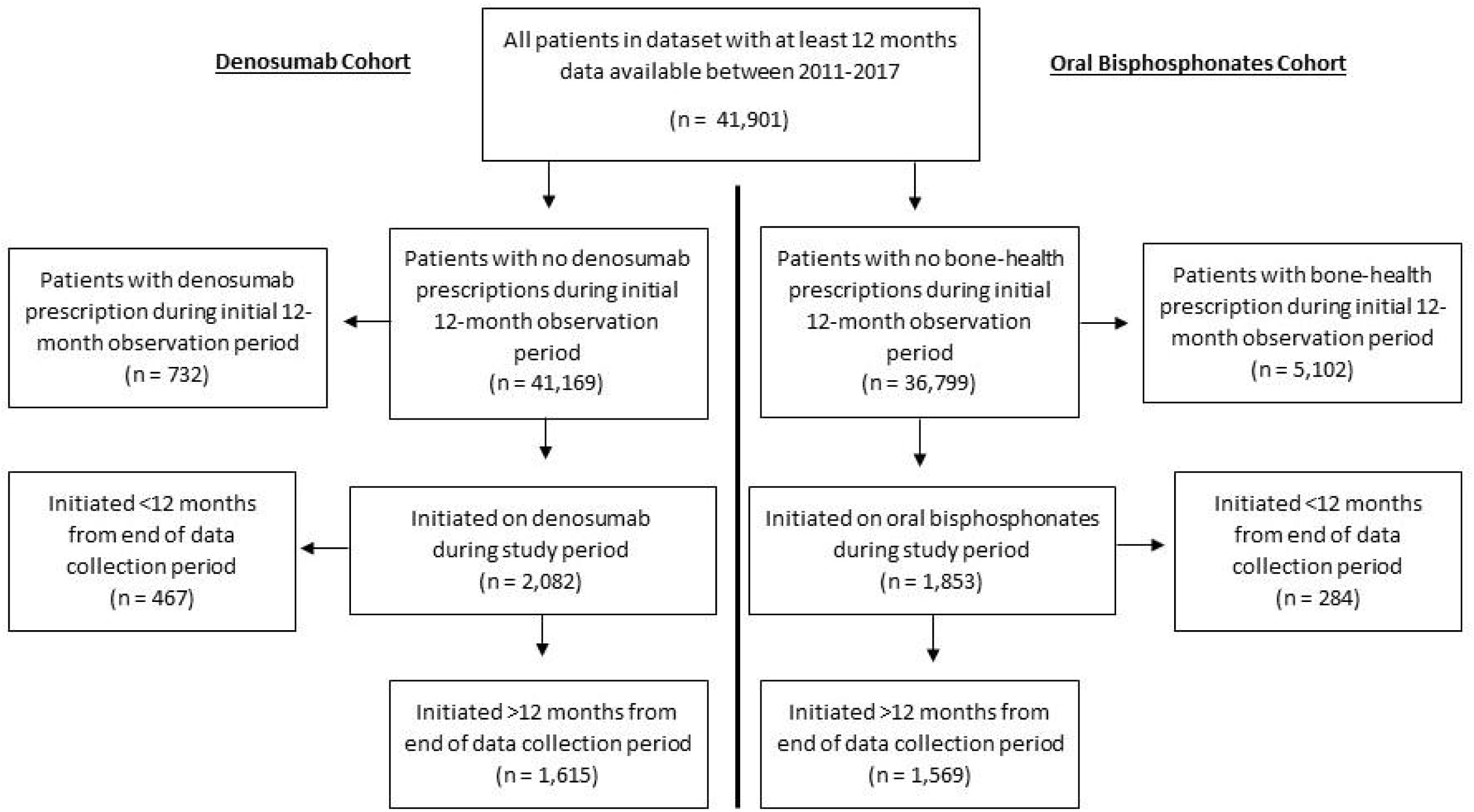
Flow-diagram of patient selection.

### Estimate of persistence

For oral bisphosphonates and denosumab, n=1,212 and n=1,146 patients respectively had at least 2 years between medication initiation and the end of data collection and did not switch to an alternative bone-health medication. Among these groups, two-year persistence was 49.4% (95% CI 46.5% to 52.2%) for bisphosphonates, and 53.8% (95% CI 50.9% to 56.8%) for denosumab.

### Factors associated with time to discontinuation of oral bisphosphonates

A total of n=1,487 patients were included in the time to discontinuation of oral bisphosphonates analysis. In the n=801 patients who discontinued bisphosphonates without switching onto another bone-health medication, mean time to discontinuation was 295 days (SD=332 days). Figure 2 shows a Kaplan Meier graph of time to discontinuation of bisphosphonates by dosing frequency. Those on monthly regimens had a higher risk of discontinuation (log-rank test, p=0.02).

**Figure 2.**
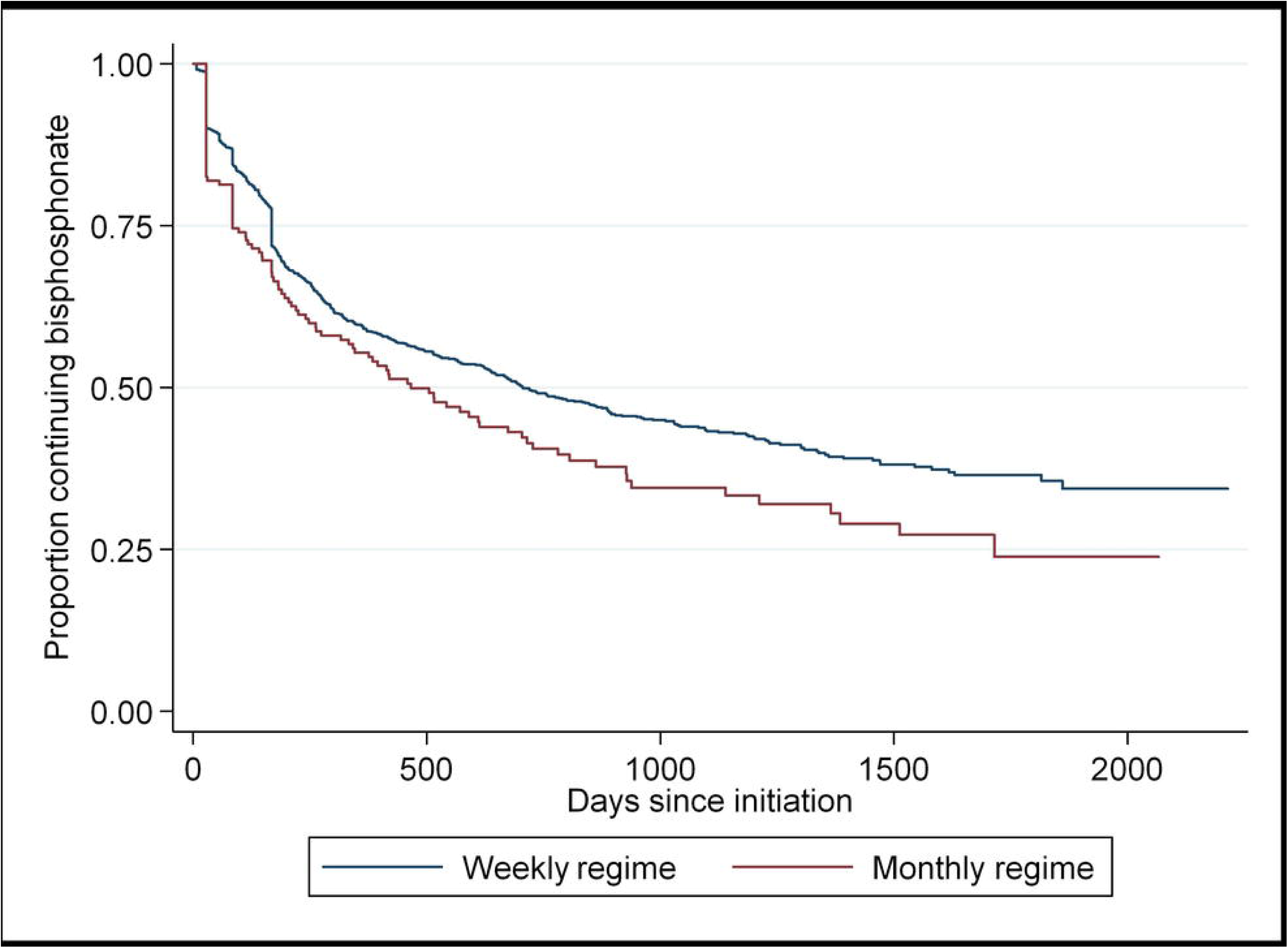
Kaplan Meier graph of time to discontinuation of bisphosphonates by dosing frequency.

On multivariable analysis (Table 2), being 80 years or older (HR=1.26, 95% CI=1.04 to 1.52, p=0.02) was associated with a higher hazard of discontinuation of oral bisphosphonates. GMS health cover (HR=0.49, 95% CI=0.36 to 0.66, p<0.01), prescription of calcium or vitamin D (HR= 0.79 95% CI= 0.66 to 0.93, p<0.01) and being on 6-10 medications rather than 0-5 medications (HR= 0.82 95% CI= 0.69 to 0.98, p=0.03) was associated with a lower hazard of discontinuation of oral bisphosphonates. The relationship between time to discontinuation and oral bisphosphonate dosing frequency did not remain statistically significant on multivariable analysis.

**Table 2.** Factors associated with time to discontinuation of oral bisphosphonates (n=1,487)

|  |  |  |  |  |  |  |  |  |
| --- | --- | --- | --- | --- | --- | --- | --- | --- |
|  |  |  | **Univariable** |  |  | **Multivariable** |  |  |
|  | **Maintained**<br>**(n=686)** | **Discontinued**<br>**(n=801)** | **HRR** | **95% CI** | **p-value** | **HRR** | **95% CI** | **p-value** |
| *Age-group at initiation:* |  |  |  |  |  |  |  |  |
| <70 years | 158 (23.0%) | 188 (23.5%) | REF |  |  |  |  |  |
| 70-79 years | 266 (38.8%) | 317 (39.6%) | 0.99 | 0.82 to 1.18 | 0.88 | 1.16 | 0.94 to 1.44 | 0.17 |
| ≥80 years | 262 (38.2%) | 296 (37.0%) | 1.01 | 0.81 to 1.25 | 0.95 | 1.26 | 1.04 to 1.52 | 0.02* |
| Sex (% female) | 527 (76.9 %) | 640 (80.0%) | 1.04 | 0.85 to 1.28 | 0.69 | 1.04 | 0.83 to 1.3 | 0.71 |
| *Health Cover:* |  |  |  |  |  |  |  |  |
| Private and Other | 98 (14.3%) | 160 (20%) | REF |  |  |  |  |  |
| GMS | 562 (81.9%) | 528 (66%) | 0.49 | 0.38 to 0.64 | <0.01* | 0.49 | 0.36 to 0.66 | <0.01 |
| DVC | 26 (3.8%) | 112 (14%) | 1.18 | 0.86 to 1.63 | 0.30 | 1.11 | 0.78 to 1.57 | 0.57 |
| Osteoporosis diagnosis | 211 (30.8%) | 229 (28.6%) | 0.90 | 0.73 to 1.09 | 0.27 | 0.86 | 0.70 to 1.05 | 0.14 |
| Fracture history | 82 (12%) | 67 (8.4%) | 0.82 | 0.64 to 1.04 | 0.11 | 0.82 | 0.64 to 1.05 | 0.11 |
| Calcium/ vitamin D prescription | 589 (85.9%) | 634 (79.2%) | 0.74 | 0.62 to 0.89 | <0.01* | 0.79 | 0.66 to 0.93 | <0.01* |
| *Number medications in previous 12-months:* |  |  |  |  |  |  |  |  |
| 0 to 5 | 151 (22%) | 252 (31.5%) | REF |  |  |  |  |  |
| 6 to 10 | 217 (31.6%) | 250 (31.2%) | 0.71 | 0.60 to 0.85 | <0.01* | 0.82 | 0.69 to 0.98 | 0.03* |
| 11 to 15 | 177 (25.8%) | 158 (19.7%) | 0.64 | 0.49 to 0.83 | <0.01* | 0.77 | 0.59 to 1.01 | 0.06 |
| > 15 | 141 (20.6%) | 141 (17.6%) | 0.70 | 0.57 to 0.85 | <0.01* | 0.83 | 0.66 to 1.05 | 0.12 |
| *Dosing frequency:* |  |  |  |  |  |  |  |  |
| Weekly | 624 (91%) | 697 (87%) | REF |  |  |  |  |  |
| Monthly | 62 (9%) | 104 (13%) | 1.28 | 1.03 to 1.59 | 0.02* | 1.21 | 0.97 to 1.50 | 0.09 |
| GMS= General Medical Services scheme<br>DVC= Doctor Visit Card<br>* P<0.05 |  |  |  |  |  |  |  |  |

### Factors associated with time to discontinuation of denosumab

A total of n=1,568 patients were included in the time to discontinuation of denosumab analysis. In the n=782 patients who discontinued denosumab without switching onto another bone-health medication, mean time to discontinuation was 401 days (SD=321 days). Figure 3 shows a Kaplan Meier graph of time to discontinuation of denosumab.

**Figure 3.**
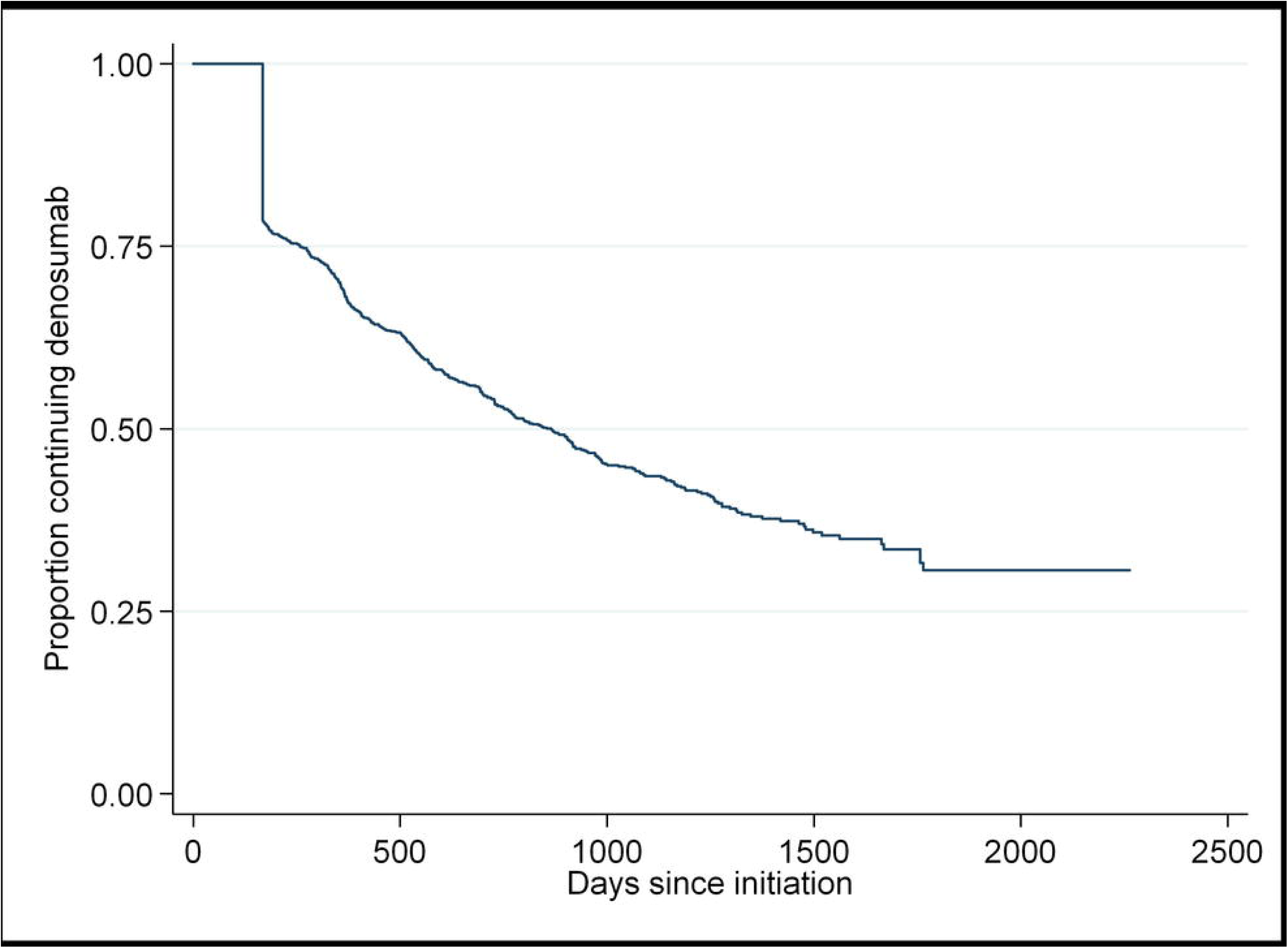
Kaplan Meier graph of time to discontinuation of denosumab.

On multivariable analysis (Table 3), no factors were found to be associated with a higher hazard of discontinuation of denosumab. GMS health cover (HR=0.71 95% CI= 0.57 to 0.89, p<0.01), and having a diagnosis of osteoporosis (HR= 0.76 95% CI= 0.69 to 0.84, p<0.01) were associated with a lower hazard of discontinuation of denosumab.

**Table 3.** Factors associated with time to discontinuation of denosumab (n= 1,568)

|  |  |  |  |  |  |  |  |  |
| --- | --- | --- | --- | --- | --- | --- | --- | --- |
|  | Maintained<br>(n=786) | Discontinued<br>(n=782) | Univariable |  |  | Multivariable |  |  |
|  |  |  | HRR | 95% CI | p-value | HRR | 95% CI | p-value |
| *Age-group at initiation:* |  |  |  |  |  |  |  |  |
| <70 years | 121 (15.4%) | 161 (20.6%) |  |  |  |  |  |  |
| 70-79 years | 291 (37.0%) | 261 (33.4%) | 0.82 | 0.71 to 0.96 | 0.01 * | 0.91 | 0.75 to 1.11 | 0.36 |
| ≥80+ years | 374 (47.6%) | 360 (46.0%) | 0.93 | 0.80 to 1.07 | 0.32 | 1.05 | 0.86 to 1.27 | 0.65 |
| Sex (% female) | 712 (90.6%) | 707 (90.4%) | 0.83 | 0.68 to 1.03 | 0.09 | 0.89 | 0.72 to 1.09 | 0.26 |
| *Health Cover:* |  |  |  |  |  |  |  |  |
| Private and Other | 122 (15.5%) | 133 (17%) | REF |  |  |  |  |  |
| GMS | 582 (74.1%) | 538 (68.8%) | 0.71 | 0.59 to 0.87 | <0.01 * | 0.71 | 0.57 to 0.89 | <0.01 * |
| DVC | 82 (10.4%) | 111 (14.2%) | 0.90 | 0.73 to 1.11 | 0.34 | 0.9 | 0.69 to 1.16 | 0.41 |
| Osteoporosis diagnosis | 402 (51.2%) | 341 (43.6%) | 0.75 | 0.68 to 0.82 | <0.01 * | 0.76 | 0.69 to 0.84 | <0.01 * |
| Fracture history | 106 (13.5%) | 77 (9.9%) | 0.80 | 0.61 to 1.06 | 0.13 | 0.83 | 0.62 to 1.11 | 0.21 |
| Calcium/ vitamin D prescription | 696 (88.6%) | 676 (86.5%) | 0.88 | 0.75 to 1.04 | 0.13 | 0.96 | 0.81 to 1.14 | 0.66 |
| *Number medications in previous 12-months:* |  |  |  |  |  |  |  |  |
| 0 to 5 | 138 (17.6%) | 149 (19.1%) | REF |  |  |  |  |  |
| 6 to 10 | 221 (28.1%) | 227 (29%) | 0.97 | 0.82 to 1.15 | 0.71 | 1.04 | 0.87 to 1.25 | 0.68 |
| 11 to 15 | 218 (27.7%) | 229 (29.3%) | 0.97 | 0.82 to 1.14 | 0.69 | 1.02 | 0.86 to 1.19 | 0.85 |
| >15 | 209 (26.6%) | 177 (22.6%) | 0.91 | 0.78 to 1.07 | 0.25 | 0.96 | 0.82 to 1.13 | 0.62 |
| Previous bone-health medication use | 335 (42.6%) | 326 (41.7%) | 0.90 | 0.81 to 1.01 | 0.07 | 0.95 | 0.85 to 1.08 | 0.45 |
| GMS= General Medical Services scheme<br>DVC= Doctor Visit Card<br>* P<0.05 |  |  |  |  |  |  |  |  |

## DISCUSSION

### Summary of findings

This study includes a large and representative cohort of older adults in the primary care setting with a long period of follow-up between 2012 and 2017. To our knowledge, it is the first estimate of persistence in bone-health medication in a general older population in the Republic of Ireland since 2009 and the wide-spread introduction of denosumab [11, 27]. Findings of sub-optimal two-year persistence (49% for oral bisphosphonates and 54% for denosumab) in the current study are in line with previous research [13, 14]. Having state-funded health cover was the only factor found to be protective against discontinuation of both medications.

### Findings in the context of previous research

For over half of patients in this study who were started on denosumab, it was the first bone-health medication they were observed to take, despite it not being recommended as a first-line treatment in most cases [6-8, 21]. This recommendation is due in part to the cost of the medication but also due to the need to pre-screen for hypocalcaemia and co-morbidities and due to complications that arise with cessation of the drug [7, 21, 28]. This pattern of prescribing reflects findings from a large primary care study in Australia where denosumab went from making up a small percentage of bone-health prescriptions in 2012 to being the most frequently prescribed in 2017 [23]. The denosumab cohort in this study included a higher proportion of female patients and more patients with a diagnosis of osteoporosis in comparison to the bisphosphonate cohort. This aligns with the strength of evidence for denosumab among women at highest risk of fracture [5, 7, 9]. The rate of contraindication to oral bisphosphonates among this group is not known, however it is unlikely to explain the rate of denosumab prescribing as first-line therapy. Due the increased popularity of the medication among GPs in recent years, further investigation of the reasoning behind these treatment decisions is warranted.

A particularly concerning finding is that only 6% of those who discontinued on denosumab were switched to an alternative bone-health medication despite this being strongly recommended by current evidence [22] For those not switched to another medication, only 55% continued taking denosumab without a gap in treatment for two-years, which is similar to findings of a recent systematic review including 16 studies from the USA, Canada and sixteen European countries [14]. Where denosumab injections are received 9-12 months apart as opposed to 6-monthly, bone turnover markers increase significantly while increases in BMD drop by over half [18, 19]. A post-hoc analysis of a randomised controlled trial of 1,001 participants, also found the rate of vertebral fractures increases five-fold on discontinuation of denosumab, quickly approaching the fracture rate observed on placebo [20]. While switching to oral bisphosphonates can protect against these changes in most patients, recently published results of a randomised controlled trial of 61 patients on longer-term therapy found that a single dose of zoledronate infusion was not sufficient to maintain benefits [21, 22]}. GPs in Australia have expressed an awareness of the quick reversal of BMD gains after stopping denosumab but also uncertainty about how and when to stop denosumab or the risks of doing so [23]. It is likely that GPs in Ireland have similar concerns and education and support in this area appears to be urgently required.

Poor persistence on oral bisphosphonates is also a concern. A 2011 meta-analysis of five studies and over 100,000 patients indicated that fracture risk increased by up to 40% with non-persistence of bisphosphonates [12]. In the oral bisphosphonate cohort in the current study, two-year persistence was estimated at 49% between 2012 and 2017, showing no improvement on older work [11, 27]. Two previous Irish studies of bisphosphonate persistence between 2005 and 2009 showed a 1-year rate of less than 50% in patients hospitalised with a fragility fracture [27] and a 2-year rate of 50% in the general older population [11]. In five studies from the USA, Canada, Hungary and Sweden that were included in a recent systematic review and that measured two-year persistence using similar treatment gaps as the current analysis, estimates ranged from 19% to 46% [13, 29-33]. While there is some clinical uncertainty about whether particular patients should be given a break or “bisphosphonate holiday” after 3-5 years to avoid increasing the risk of adverse events, this should not influence persistence after only two years on medication [15-17, 23]. Furthermore, this cohort would be considered at relatively higher risk of fragility fracture as they have an average age of 77, a fracture history prevalence of 10% and a diagnosis of osteoporosis in 30% of the group. Guidelines suggest that among patients at high fracture risk, alendronic acid may be safely continued for up to 10 years and risedronate for up to seven years [6]. It should be noted that our estimate of persistence could be optimistic as we used a conservative acceptable treatment gap of 90 days and excluded switchers onto alternative medications [13, 34]. In addition, in contrast to previous research, our study did not find more frequent dosing regimens of oral bisphosphonates to be associated with discontinuation [27, 30-32, 35]. In fact, on univariable analysis, monthly regimens had a higher Hazard Ratio than weekly formulations. This may reflect monthly formulations being targeted towards patients likely to have challenges persisting to the prescribed regimen.

Having state-funded health cover (GMS) was the only factor found to be protective against discontinuation of both oral bisphosphonates and denosumab in this study. This relationship remained strong even after adjusting for age. This is important, as the GMS scheme in Ireland is means-tested but a higher income threshold applies to those aged 70 and over [26]. For this reason, 50-55% patients in this study aged under 70 years were covered by DVC/GMS in comparison to 90% of patients 70 years and older. Such patients who have free access to GPs, practice nurses and medications may be more likely to return for repeat prescriptions, support and administration of medication (in the case of denosumab). Older age group (80 years and older) showed some association with discontinuation of oral bisphosphonates on multivariable analysis, independent of health cover, but this was not observed in the denosumab cohort. In previous literature, age has shown an inconsistent relationship with persistence of these medications with both the oldest (>75 years) and youngest (<65 years) most likely to discontinue [13, 34, 36, 37]. This may be related to an increased likelihood of adverse effects at older ages or patients or physicians not prioritising treatment of fracture risk in younger patients [34]. As this study included only patients who were aged over 65 by 2017, this could have resulted in higher persistence overall.

In our study, prescription of calcium or vitamin D was associated with a lower hazard of discontinuation of oral bisphosphonates and having a recorded diagnosis of osteoporosis was associated with a lower hazard of discontinuation of denosumab. This could potentially be explained by ongoing osteoporosis management being reflective of the patient and physician prioritising the need for therapy, which is suggested to be an important determinant of adherence to these medications [34]. Prior BMD testing, using other drugs for osteoporosis and calcium or vitamin D supplementation have been associated with better persistence in previous research [13, 35, 38]. In contrast to several other studies however, a history of fragility fracture was not found to be associated with improved persistence in our analysis [13, 38]. This is surprising, as one would expect a fracture experience to highlight the need for treatment and improve the management pathway. Studies in Australia and Canada have found that for secondary fracture prevention, while specialist-led programmes can facilitate better initiation of therapy, primary care physician follow-up is as effective at improving persistence [39, 40]. This suggests that GPs could be supported to provide long-term management of osteoporosis in patients with fragility fracture but that once-off reviews with geriatricians could be beneficial. This requires further investigation in the Irish setting.

### Strengths and limitations

A strength of this study is that we ascertained prescribing from multiple sources (i.e. GP prescription records and hospitalisation discharge summaries). We included a washout period to look at those newly initiated on medication as patients have been found to be more persistent if evaluated from their first exposure to osteoporosis therapy [41]. Using routinely collected data, we were unable to assess reasons for discontinuation of medication that may have been clinically appropriate and do not know if it resulted from a risk-benefit discussion with patients. We were also unable to determine if patients received prescriptions/ treatment (including denosumb or bisphosphonate infusion) solely from outpatient appointments with hospital-based specialists or during hospital admissions. Regardless, the very high rate of non-persistence to denosumab without observed replacement by bisphosphonates or other therapies within the primary care setting is a major concern. As data related to prescribing, it is not possible to determine whether prescriptions were dispensed, or if patients took their medication as prescribed. This may have resulted in an optimistic persistence rate. Finally, it is unknown, whether those “initiated” could have been finishing a “bisphosphonate holiday” or those discontinuing could have re-initiated later on. Discontinuing denosumab however has significant risks in the short-term and so looking for delayed re-initiations was not an objective of our analysis.

### Clinical implications

Non-persistence/ adherence to osteoporosis medications is wasteful and can pose significant patient risks, especially in the case of denosumab treatment. Further research is required in the Irish primary care setting to explore the reasons for prescribing choices and patterns and to evaluate interventions targeted at both patients and physicians. A recent systematic review found that multi-component education programmes that included patients in the decision-making process around osteoporosis treatment and specific regimens improved medication persistence [42]. A 2012 Irish analysis suggested that investing €120 annually per patient into interventions would remain cost-effective if they improved adherence and persistence to osteoporosis medication by just 10% [11]. This warrants further testing.

## CONCLUSION

This study has identified a number of areas where fracture preventive prescribing among older adults in primary care could be improved. This includes the common use of denosumab as a first-line treatment, sub-optimal rates of persistence with bisphosphonates and denosumab at two years and low rates of switching to other preventative treatments among those stopping denosumab. Free access to primary care services and medications may facilitate persistence, however other interventions targeting patients and prescribing in primary care to optimise prescribing warrant evaluation.

## Data Availability

No additional data available.

## DECLARATIONS

### Availability of data and material

No additional data available.

### Authors’ contributions

MEW, TF, and FM conceptualised the study. MEW and FM performed the analysis. MEW drafted the initial manuscript. All authors critically appraised and edited the manuscript. FM is the guarantor. All authors read and approved the final manuscript.

### Ethics approval

Ethical approval was obtained from the Irish College of General Practitioners.

### Consent to participate

This study involved the analysis of anonymous routinely collected data. No individual patient consent was obtained.

