## Appendix for "Persistence with oral bisphosphonates and denosumab among older adults in primary care in Ireland"

Journal: Osteoporosis International

Authors: Mary E. Walsh^1^ , Tom Fahey^1^ , Frank Moriarty^1,2^

Affiliations: ^1^HRB Centre for Primary Care Research, Dept. of General Practice, Royal College of Surgeons in Ireland, Dublin, Ireland; ^2^School of Pharmacy and Biomolecular Sciences, Royal College of Surgeons in Ireland, Dublin, Ireland

| **Online Resource 1. Prescription and condition definitions** | | |
| --- | --- | --- |
| **Medication/ Diagnosis** | **Codes** | **Free-text search terms** |
| **Bone-health medication:** |  |  |
| Oral Bisphosphonates | ATC: M05BA01 M05BA02 M05BA03 M05BA04 M05BA05 M05BA06 M05BA07 M05BB | aclasta actonel alendromax alendronic aredia binosto bonapenya bonasol bondenza bondronat bonefos bonefurbit bonviva clasteon destara didronel pamidronate fosalen fosamax fostepor fostolin iasibon ibandronic ibandronate kefort loron optinate osbonelle osteomel ridate risedronate riseseus risonate risontel romax teboneva tevanate zerlinda adrovance alendronate didronel fosavance optinate ridate vantavo etidronic etidronate clodronic clodronate pamidronic tiludronic tiludronate alendronaate aledronic alenronic risedronic |
| Denosumab | ATC: M05BX04 | denosumab prolia xgeva |
| Bisphosphonate Infusions | ATC: M05BA08 | zoledronic zometa zoledronate |
| Raloxifene | ATC: G03XC01 | raloxiep raloxifene |
| Parathyroid hormone | ATC: H05AA | parathyroid forsteo movymia natpar preotact terrosa tetridar teriparatide |
| Strontium | ATC: M05BX03  ATC: M05BX53 | osseor protelos strontium |
| Calcitonin | ATC: H05BA | calsynar forcaltonin miacalcic miakaril ostulex calcitonin elcatonin |
| **Other Exposure Variables:** |  |  |
| Osteoporosis | ICD-10-AM: M80-M82  ICPC 2: L95 | osteop paget |
| Fragility Fracture | ICD-10: M48.4 M48.5 S22.0 S22.1 S32.0 S42 S52 S7  ICPC2: L72 L75 L76 | colles "wrist fracture" "distal radius fracture" "compression fracture" "fatigue fracture""neck of femur" "hip #" "nof" "osteoporosis with pathological fracture" "fracture of lower end of radius" "fracture of humerus" "vertebral collapse" "hip fracture" "femoral fracture" |
| Prescription of calcium and/ or vitamin D | ATC: A12A A11CC A11CB | actonel altavita "at 10" bellcalcid bocatriol cacit cadelius calcichew calciforte calcijex calciup cal-d-vita calfovit caltrate calvidin crampex desunin everose fultium-d3 "halibut liver oil" ideos kalcipos-d "one-alpha" osteocur osteofos ostram osvaren rocaltrol sandocal sapvit-d3 silkis teboneva thorens calcium ergocalciferol dihydrotachysterol alfacalcidol calcitriol colecalciferol calcifediol "vitamin a and d" "vitamin d" cholecalciferol calciche calciferol cholecalciferol calcihew calcidol alfacalcidiol calcitrol cholecalcierol |
